# Impact of B.1.1.7 variant mutations on antibody recognition of linear SARS-CoV-2 epitopes

**DOI:** 10.1101/2021.01.06.20248960

**Authors:** Winston A. Haynes, Kathy Kamath, Carolina Lucas, John Shon, Akiko Iwasaki

## Abstract

In 579 COVID patients’ samples collected between March and July of 2020, we examined the effects of non-synonymous mutations harbored by the circulating B.1.1.7 strain on linear antibody epitope signal for spike glycoprotein and nucleoprotein. At the antigen level, the mutations only substantially reduced signal in 0.5% of the population. Although some epitope mutations reduce measured signal in up to 6% of the population, these are not the dominant epitopes for their antigens. Given dominant epitope patterns observed, our data suggest that the mutations would not result in immune evasion of linear epitopes for a large majority of these COVID patients.

## Introduction

The B.1.1.7 strain circulating within the UK has raised public concerns about potential for reinfection and vaccine efficacy due to possible evasion from antibody recognition. The non-synonymous mutations of the B.1.1.7 strain of SARS-CoV-2 are characterized by 2 deletions and 6 mutations in spike glycoprotein, an early stop codon and 2 mutations in non-structural protein 8, and 2 mutations in nucleoprotein^1^. Whether these mutations render pre-existing antibodies ineffective has become a public concern as it could result in reinfection or loss of efficacy against vaccination. Here, we apply a high throughput, random bacterial peptide display technology called serum epitope repertoire analysis (SERA)^2,3^ that enables assessment of SARS-CoV-2 seropositivity and high-resolution mapping of epitopes across any arbitrary proteome. In this case, we examine wild-type SARS-CoV-2 and the B.1.1.7 variant, to interrogate whether the mutations that are found in the B.1.1.7 variant renders existing antibodies incapable of binding to their viral targets.

## Results

In order to determine whether the non-synonymous mutations in the B.1.1.7 strain block binding by antibodies generated during natural infection with the existing SARS-CoV-2, we performed SERA^2,3^ on 579 COVID-19 patients (all collected prior to the emergence of B.1.1.7), which we thoroughly described in Haynes, Kamath, Bozekowski, *et al*^4^. From the SERA analysis with a bacterial display library presenting random 12mer amino acid sequences, we acquired a set of 12mers for each patient which represent the patient’s antibody epitope binding specificities. Using protein based immunome wide association studies (PIWAS)^5^, we tiled the COVID-19 patient SERA data *in silico* against the B.1.1.7 proteome and the wild type SARS-CoV-2 proteome.

### Spike glycoprotein

Spike glycoprotein in the B.1.1.7 contains 2 deletions and 6 point mutations. First, we noted that the regions harboring mutations generally had no direct overlap with the epitope hotspots we have previously described^4^ [Figure 1A]. The epitope mutations with the most dramatic reduction in outliers (defined as epitope score > 99^th^ percentile of the pre-pandemic controls) were del144, N501Y, A570D, and P681H. Mutational changes resulted in only 2 individuals (0.3% of the population) having a dramatic reduction in PIWAS antigen scores, which reflects the peak epitope signal along the entire antigen.

**Figure 1.**
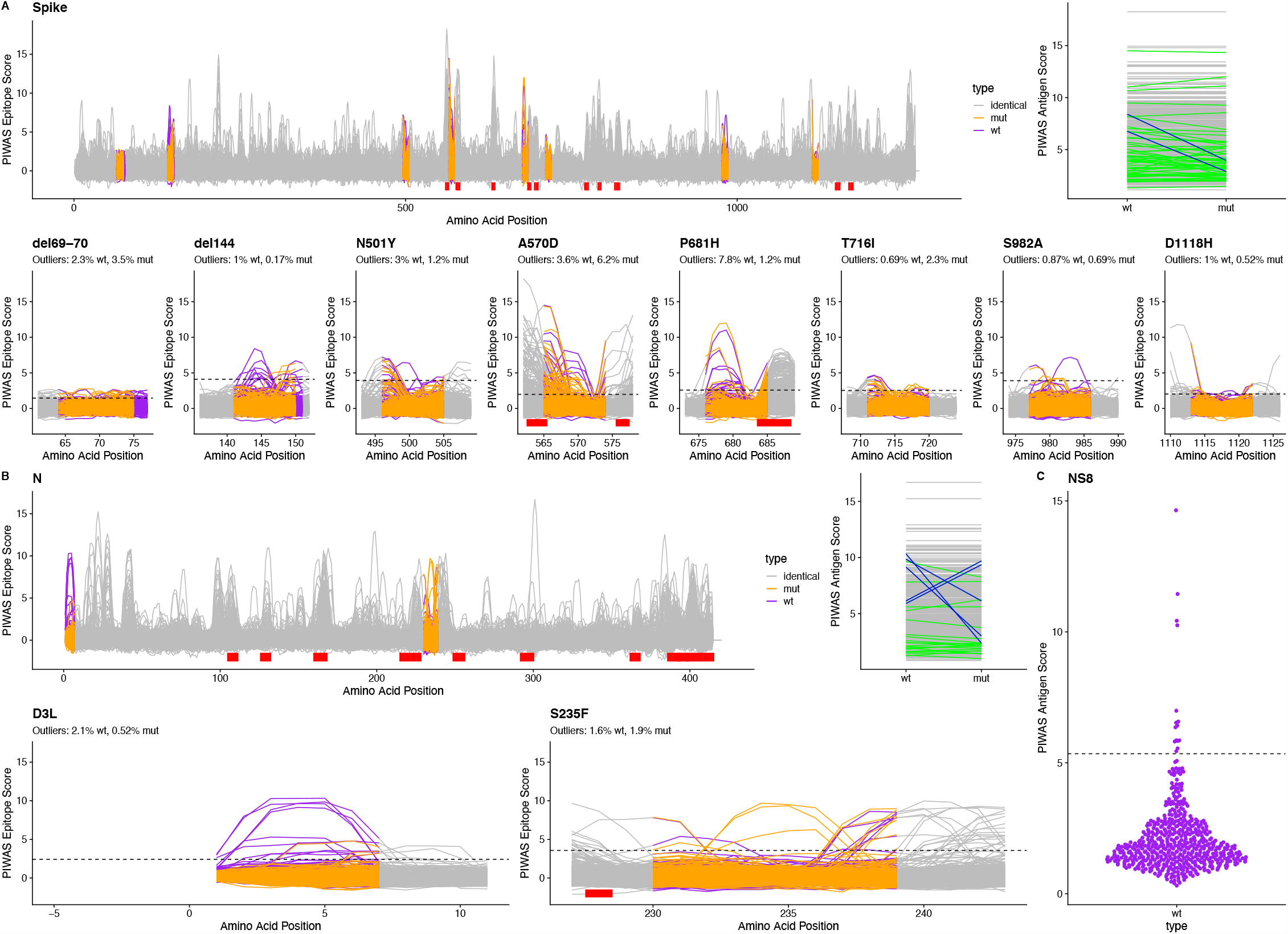
Linear epitope signal in COVID-19 patients for B.1.1.7 SARS-CoV-2. We used PIWAS analysis to examine linear epitope signal against B.1.1.7 SARS-CoV-2 in 579 COVID patients. For (A) spike glycoprotein and (B) nucleoprotein, we performed PIWAS tiling against both wild type (purple) and B.1.1.7 (orange) SARS-CoV-2. Regions outside of the variant region are tiled against the wild-type protein and are marked as identical (grey). Dominant SARS-CoV-2 epitopes identified in Haynes, Kamath, Bozekowski, *et al*^4^. are shown with red bars. PIWAS antigen score highlights individuals where the signal changed (green) or decreased by at least 3 (blue) as a result of the strain variants. For each epitope, we show the percentage of outliers (dashed line, PIWAS score > 99^th^ percentile of the pre-pandemic control population) for the wild type and B.1.1.7 mutant. (C) We show non-structural protein 8 PIWAS antigen scores for the wild type SARS-CoV-2 strain relative to the 99^th^ percentile in the pre-pandemic control population.

### Nucleoprotein

The B.1.1.7 strain harbors 2 point mutations: D3L and S235F. At the epitope level, the D3L mutation reduces PIWAS signal in 1.5% of COVID patients (outliers reduce from 2.1% in wild type to 0.5% in mutant) [Figure 1B]. In 3 of these patients (0.5% of the population), that alteration represents the strongest antigen signal and substantially reduces the PIWAS antigen score. Surprisingly, the S235F substantially increases epitope and antigen scores in 2 individuals. At a population level, the net result is a neutral effect on predicted nucleoprotein antigen signal.

### Non-structural protein 8 (NS8)

PIWAS antigen scores for 2.5% of the COVID patients exceeded the 99^th^ percentile of the pre-pandemic controls [Figure 1C].

## Discussion

We rapidly characterized the predicted changes in epitope response as a result of the B.1.1.7 strain of SARS-CoV-2 using the randomized bacterial display of SERA coupled with proteome analysis using PIWAS. Through this combination, we are able to computationally analyze signal against new proteomes without rescreening any samples. In COVID patients, we observe limited changes in epitope signal on the B.1.1.7 strain compared to the original strain. Although the early stop codon in NS8 resulted in the most decreased signals, antibodies to NS8 are unlikely to play a role in a protective response. Given the suggested role of NS8 in severe disease^6^, this early stop codon might have a positive effect on disease outcome.

Our data suggest that the mutations seen in the B.1.1.7 strain of SARS-CoV-2 would not result in loss of dominant antibody responses to linear spike glycoprotein and nucleoprotein epitopes in the vast majority of our cohort’s COVID patients. Since PIWAS detects linear epitope signals, further assays are required to examine structural evasion of the immune response. In particular, P681H is immediately upstream of the furin cleavage site, which is a hotspot for epitope signal.

Given that our data does not yet include SARS-CoV-2 vaccinated samples, our conclusions beyond natural infection are speculative. Since the mRNA vaccines include intact spike sequences^7,8^ resembling those which are circulating in naturally infected individuals, and naturally infected individuals exhibit a somewhat durable immune response^9^, we have no evidence to suggest that the current vaccines won’t be effective against B.1.1.7.

## Methods

### Serum epitope repertoire analysis

Both a detailed description of the SERA assay^2^ and the screening of these specific samples^4^ have been previously published. As described in Haynes, Kamath, Bozekowski, *et al*: “For this study, serum or plasma was incubated with a fully random 12-mer bacterial display peptide library (1×10^10^ diversity, 10-fold oversampled) at a 1:25 dilution in a 96-well, deep well plate format. Antibody-bound bacterial clones were selected with 50 µL Protein A/G Sera-Mag SpeedBeads (GE Life Sciences, cat#17152104010350) (IgG) or by incubation with a biotinylated anti-human IgM antibody (Jackson ImmunoResearch, cat# 709-066-073) final assay dilution 1:100, followed by a second incubation with 50 ul Dynabead MyOne Streptavidin T1 conjugated magnetic beads (IgM) (Thermo-Fisher 65602). The selected bacterial pools were resuspended in growth media and incubated at 37°C shaking overnight at 300 RPM to propagate the bacteria. Plasmid purification, PCR amplification of peptide encoding DNA, barcoding with well-specific indices was performed as described. Samples were normalized to a final concentration of 4nM for each pool and run on the Illumina NextSeq500.”^4^

### PIWAS analysis

To identify linear epitope signals, we applied the previously published PIWAS method^5^ against the Uniprot reference SARS-CoV-2 proteome (UP000464024)^10^ and the B.1.1.7 variant described in Rambaut, *et al*.^1^. The PIWAS analysis was run on the IgG SERA samples described in Haynes, Kamath, Bozekowski, *et al*.^4^: a single sample per COVID-19 patient (for a total of 579 patients) versus 497 discovery pre-pandemic controls, and the 1500 validation controls used as the normalization cohort. Additional parameters include: a smoothing window size of 5 5mers and 5 6mers; z-score normalization of kmer enrichments; maximum peak value; and generation of epitope level tiling data.

### Variant analysis

We compared PIWAS epitope tiling data from spike glycoprotein, nucleoprotein, and non-structural protein 8 in the wild type and B.1.1.7 variant of SARS-CoV-2 for each subject. At the antigen level, peak wild-type vs. peak mutant PIWAS values were compared to identify individuals with decreased epitope signal at the antigen level. We included motifs from Haynes, Kamath, Bozekowksi, *et al*. ^4^ that mapped linearly to the protein sequences to highlight dominant epitopes. We compared PIWAS antigen scores for these same proteins and variants.

## Data Availability

Data available upon request.

## Acknowledgements

We would like to thank the entire team at Serimmune, collaborators, and patients who have made this study feasible. A.I. is an investigator of the Howard Hughes Medical Institute.

